# Logistic Approach to COVID - 19 Epidemic Evolution in Brazil

**DOI:** 10.1101/2020.06.22.20135921

**Authors:** Altair Souza de Assis, Vinicius Werneck de Carvalho

**Affiliations:** Private Investigator, Niterói, RJ, Brasil

**Keywords:** New Coronavirus, COVID - 19, Dynamic Models, Logistic model, Predator – Prey model, Dynastic model, Contamination Inflexion. Contamination Saturation – Plateau Regime, Social Isolation

## Abstract

We study in this work the temporal evolution of local and global contaminated population by coronavirus. We access those information analytically and numerically using a logistic model. It is shown, using diferent data from The Brazilian Ministry of Health (MS), The World Health Organization - WHO, and The Niteroi Health Foundation (FMS), the contaminated population ramping-up curves, the population inflection, the population saturation - plateau regime, and also the time related to these population evolution regimes. Based on the simulations, approaches are proposed at this more advanced phase of the pandemic, which might generate effectiveness at the actions of society in general, in a way that those actions could generate effective and efficient results, and this means a more organized war against this pandemic, a better way to induce the economy resumption, and also to create a more intense public awareness on the contamination hubs and surges that may emerge due to the reduction of social isolation.

## Introduction

By May 21, 18,894 deaths were registered in Brazil, 293,357 confirmed cases of contamination and 116,683 recovered from COVID −19. On June 11, less than 1 month later, according to the National Council of Secretaries (Conass) and the Ministry of Health(MS), there are already 40,919 deaths, 802,828 confirmed cases, and 345,595 recovered and 465,740 deaths. Today, june 22, according to the World Health Organization (www.who.int) the world number is 8,860,331 contaminated, and to Brazil according to MS (covid.saude.gov.br) it is 1,085,038 contaminated, 50,617 deaths, 516.3 contaminated per 100,000 ihabitants, mortality 24.1 per 100,000 inhabitants, 641 new cases, and letality 4.7%.

The important question to ask ourselves is: what is the real formation law for the temporal evolution function of the contamination density to ramp-up, to achieve the plateau regima and eventually to ramp-down? How to study access this function in order to take efficient and effective measures to guarantee the maximum action effectiveness to face this world public health crisis?

Numerous articles have been written showing predictions about contamination by the corona virus, in scientific journals or on news sites (1 − 5). Many of those predictions did not materialize, some overestimated and others underestimated the related numbers and functions, some others came closer to the real curve and figures, depending on the complexity of the model used or the epidemiological knowledge of the subject. However, the numerology in this coronavirus scenario is still a problem open to investigation and if any additional light is shed on this topic that everyone is interested in, it can still save lives and reduce the suffering of many families. It is clear that the world needs to be highly effective in coping with this health and later economic crisis.

Analyzing the recent data from the Brazilian Ministry of Health (MS) and the “World Health Organization (WHO)” (6), it is very clear that, in the absence of an effective vaccine to contain the COVID-19, social isolation is still the best weapon to face this pandemic surge, even in its most advanced stage. Studies show that social isolation is really the most effective measure to contain the advance and possible new contamination hubs (7), as now in China and in other parts of the globe. The social isolation tis effective when done in an organized way, considering all the social parameters involved as economic, psychological, and other relevant ones.

Intelligent control can unlock the economy and still preserving lives at the same time. Is there, then, a mathematical foundation in this scenario to ensure that social isolation is in fact a good decision to avoid a high number of unnecessary deaths, even after saturation or peak? Does this model give indications of how social isolation can become less rigid, but without a severe loss of lives? In this work, an logistic analytical model and numerical simulations are presented, simple to understand, which will show the importance of social isolation, until the viral field dissipates to a “noise level”. This dissipation can be induced by a vaccine or by other related means, therefor avoiding contamination surges and hubs resulting from the relaxation of the containment measures which could cause new waves of inevitable deaths (second wave) (8).

## Methodology

### Mathematical Models for Population Dynamics

#### 1. Exponential Growth

Let *N* (*t*) be the population of a given specie (viruses, bacteria’s, contaminated people, or any other type) over time. The most elementary hypothesis regarding the variation of the population *N* (*t*) over time is that its rate of change is proportional to its own value at time t, that is (9),

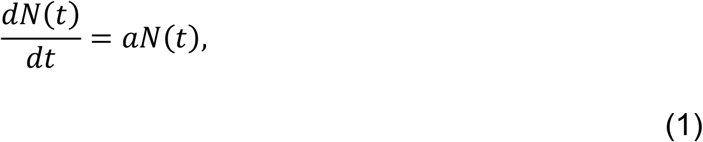

where the proportionality constant a is called the base rate for population growth or decline, depending on its sign, since it can be positive or negative, and t is time (hours, days, weeks, months, or other convenient time unit). The choice of the convenient time unit depends on the problem under consideration. Equation (1) can only be solved completely if we know the population value at *t* =0(when the timer is turned on), that is,

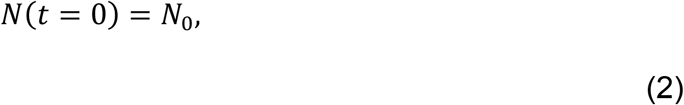

Where *N*_0_ is a problem known/given value. For this reason the system of equations (1) and (2) is called the initial value problem. Note that the choice of *t* =0 is conventional, we can choose *t* =0 at any position on the time scale that would be convenient for our problem, as long as the population is known at this time. Therefore, as said above, the population at *t* =0, is a known information and has the value *N*_0_, which is nothing less than the initial value of the number of elements in the study: people, viruses, bacterias, animals, contaminated people, and so on. Assuming that we are studying the growth and not the death of the target population, that is, considering a positive, it is obtained as a solution to the problem (by integrating equation 10) (9),

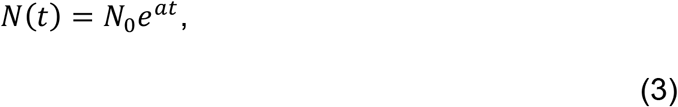

Where *e*=2,718…is the basis for the Neperian logarithms, created by the Scottish mathematician John Napier. Equation (3) from the proposed model, predicts that the population will grow exponentially over time without restriction. This is exactly the increase in the number of contaminants that we have seen in the case of Brazil and the world at the initial course of the pandemic, with a sign of saturation in some countries, and decrease in others, and an inflection of the curve in others. This exponential model does not serve to model situations where *N*(*t*) is already at the inflection phase, or saturation.

The exponential growth occurs in the initial phase of contamination, but also for longer times (as we see in the case of Brazil), when social isolation measures are not respected or not well managed and the number of contaminated people in contact with the healthy population is very large, and therefore generating a contamination rate much higher than expected, and worse without any reducing factor for the curve to stabilize. However, this type of growth does not occur indefinitely for obvious reasons, at some point it will saturate in a small or large time interval, depending on the load capacity of the system. Even considering the population of the country as a whole at some point the curve saturates, but in this case the number of contaminants would be astronomical - a large percentage of the system. The system tends to stabilize, because in a population of human beings or animals there is always a lack of food for everyone, for example, generating conflicts, wars, and diseases, so growth would start to decline, would be limited by factors not considered in the simple ideal exponential model. In any case, the number of contaminated people will eventually stabilize (with several deaths of course) by the so-called herd immunity effect (10).

Any population system has a carrying capacity, that is, any pandemic ends at some point, an extinction of that pandemic load depends on factors such as possible harmful mutations in the virus that affect its ability to reproduce and / or infect more people [in fact, recent studies by Hyeryun Choe of the Department of Immunology and Microbiology at Scripts Research, Florida Campus, noted that there were changes in the number of viruses increased in the number of “spikes”(Peplômeros – in general glycoproteins and lipids) that are actually true pointed in its crown (surface of the viral envelope of the viral separations), this allows in a greater deal the connection of viruses with the invaded cells], vaccines, herd immunity, and the reduction of the virus capacity to reproduce itself.

For short periods of time, or under extremely abundant resources, the exponential model presented works well, which is not the case in general. To study populations with growth restrictions, equation (1) is no longer valid. However, there is another model that can be used to this type of scenario called logistic model. In this model populations have regulations that depend on their own density, so it is necessary to adjust the previous model to take into account the density dependence in the evolution of the system (11).

Basically, any type of resource important for the survival of species is able to act as a system limiter, then generating competition between species in the system. Verhulst (Pierre François Verhulst - 1838) (11), derived the logistic equation to study the self - limiting growth of a population. In the Verhust model, the reproduction rate is proportional to both the existing population and the amount of available resources, everything else is considered constant (the parameters of the problem can be estimated by non-linear regression). In this way of looking at the problem, the population tends asymptotically to a saturation value (carrying capacity of the system).

The logistic function (SIGMOIDE) which is the solution to the logistic equation, is the inverse to the natural LOGIT function, it can also be used to convert the logarithm of chances into a probability. The logistic function is the solution of a first order non-linear differential equation (11, 12, 19). In this new model, populations have regulations that depend on their own densities, so it is necessary to adjust the previous model to take into account the dependence of density on the evolution of the system. This dependence can be better understood by the figure below, which shows how the population increase depletes the natural resources of the ecosystem in question. For the containment of the virus this is very good, because if little by little it has no one to infect the pandemic loses its strength.

#### 2. Logistic Growth

In the logistic model, it is considered that the rate of temporal growth of the population depends on this same population at the time of the measurement (that is, the real population is considered, with the reduction induced by the growth restrictions, at time t). The equation that models this possible population scenario is called, as seen above, Verhulst’s equation. Equation (1) can now be written as (12, 13, 14):

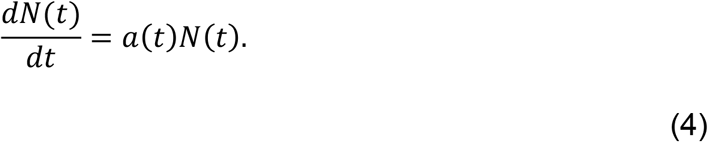

The term *a* (*t*), in general, is a complicated function of t, in the case of more complex population dynamics, from the dynamic point of view. If we consider *a*(*t*)= *a*(constant). Equation (4) is reduced to the case of exponential growth previously studied.

In the logistic model, the population control function *a*(*t*) is considered as follows (law of variation): *a*(*t*)= *a* − *bN*(*t*), with a constant. This is still a simplified model of a(t), it is considered that population growth, limited by the term *bN*(*t*), is no longer uniformly increasing over time. However, an inflection in the curve is expected at some time in the course of the evolution of the system (that is, as *bN*(*t*) grows in relation to the term a). Thus equation (4) can be written as:

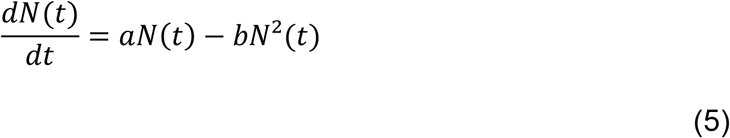

In equation (5), the initial growth is free and dominated by the first term *aNt*. The rate a represents the proportional increase in population *N*(*t*)over time, in a conveniently chosen time unit. In our case t will be measured in months. As the population grows over time, the second term, −*bN*^2^(*t*), also grows, as members of the *N*(*t*) population strat to interfere with each other, competing for critical system resources, such as food, medicine, and territory, or people to infect in our case. This 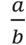 antagonistic effect is called “bottleneck”, it is modeled by the value of the population saturation parameter. The competition in the population decreases the combined rate of growth, until the value of *N*(*t*) stops growing over time, thus reaching the so-called maturity of the population *N*_*sat*_ (called the saturation value of *N*(*t*), where *N*(*t*) no longer grows over time).

This ordinary non-linear differential equation is, fortunately, the famous Ricatti equation with solution easily to be obtained. It is given by (12):

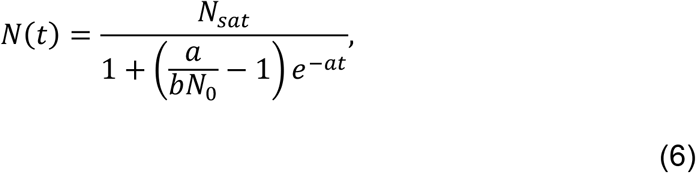

Where *N*_0_is the value of *N*(*t*) at *t* =0.

It can be seen that equation (6) is slightly different from the exponential evolution (3). Equation (6) clearly indicates the need to avoid new contagions so that an inflection point occurs in *N*(*t*)and a therefore a saturation level. After that, we expect the subsequent time decrease in the contaminated population. In order to have that, the model shows that the most effective way to contain a high rate of contamination of the healthy population and obtain a quick inflection is via quarantine/social isolation, strict epidemiological control, and produce a vaccine as soon as possible. This is to avoiding overloading country’s health system. The b parameter clearly points to the direction of a real social isolation. Effective quarantine /social isolation and strict personal protection for healthy, sick, or asymptomatic people seems to be a good strategic decision, supported mathematically by the above model.

Smaller 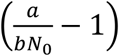, faster the system saturates and fewer people die. This occurs if *a* ≪ *bN*(*t*), where *b* is a parameter that “models” the lack of healthy people (in the sense of not having had the virus yet) to be contaminated, and the virus’s inability to reproduce effectively (15,16). At this current phase of relaxation of social isolation measures, the two concepts presented below are important.

In the case of the coronavirus, there are several doubts on the possible recontamination processes, distance of the virus’s effective kinetic propagation through the circulating air (is there any air runaway virus/droplet’s due to some runaway air molecules?), virus life time on surfaces (fomites), and other related subjects. When there is doubt about an unknown thread, we should not neglect the risk, one should always use the well-known “precautionary principle”. The precautionary principle states that the burden of proof of potentially dangerous actions by industry or the government rests on ensuring safety and that, when there are threats of serious harm, scientific uncertainty must be resolved in favor of prevention (17). For those who are professional or not, who need to expose themselves to a known or even unknown risk, they can also use the “ALARA” principle that takes into account the exposure time, the distance from the source, and the shielding. Would its use be different for exposure to global biological risks (18)?

In equation (6), the growth speed decreases with time as *bNt*grows over time, that is, at the beginning, we have *a* ≫ *bN*(*t*)and the growth is of exponential type. As *bN*^2^(*t*)increases and *a* − *bN*(*t*)decreases, the exponential starts to be attenuated until the curve undergoes an inflection, which will occur at *N*(*t*=*t*_*inf*_). At this point, where *N* (*t*)= *N*(*t* = *t*_*inf*_), it is the exact instant where the curve that models *Nt*undergoes an inflection, from there it grows with a tendency to saturate at *t*_*sat*_, where *t*_*sat*_ is the exact moment when the curve starts the saturation process. From the logistic model it is known that *N*_*inf*_ = *N*_*sat*_/2.

Mathematically, it is not very trick to find the inflection points or critical points of a curve, it is enough to cancel the second derivative of the function that describes this curve. However, if it does not cancel out and it is positive, the curve has a positive concavity (facing upwards) and if it is negative it has a negative concavity (facing downwards). As well known inflection point separates the positive from the negative concavity in a curve.

From the inflection point, the population density solution, solution curve of the logistic equation, inverts its position (or sign, from positive to negative), until eventually saturates at the population density *N*(*t* = *t*_*sat*_) = *N*_*sat*_ (*t*) = *N*_*sat*_(*t* = ∞). The saturation density can be obtained by the equality *a* = *bN*(*t* = *t*_*sat*_), this implies that equation (5) has 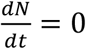, which means that *N*(*t*)is constant for any subsequent *t*.

After saturation, the curve can only remain at this level or decrease over time, which would be the natural trend under normal conditions, and it is what is expected from the number of people infected by the coronavirus. Since *b*=*a*/*N*_*sat*_,, we have that the temporal evolution of the contaminated population is given by (logistic equation):

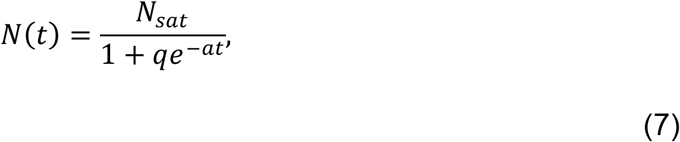

Where 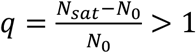, since *N*_*sat*_>*N*_0_for any time t. To know the exact saturation time, just make *N*(*t*) = *N*(*t*_*sat*_) = *N*_*sat*_ in (7), that is 1 + *qe*^− *at*^ = 1 (*t* = *t*_*sat*_). Thus, 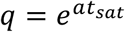, implying that In(*q*) = *at*_*sat*_, *or* 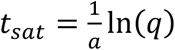. Logo the saturation time, in terms of the contaminated population, after replacing q by tits original value, is:

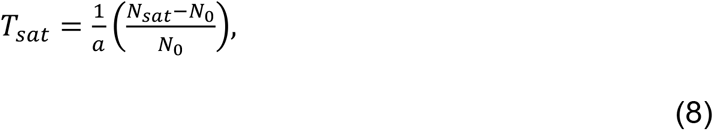

As can be seen, the logistic equation (7) contains four parameters, namely: *N*_0_, *a, b, N*_*sat*_. The initial density *N*_0_ as already seen is a known information of the problem, the rest of the parameters can be determined precisely by regression analysis (19, 20). The logistical approach is not linear, so care must be taken when choosing the starting parameters. However, there is a simpler way to obtain those parameters, to write the logistic equation with all the parameters explicit, and for that it is enough to know three equidistant values of N(t), which in this study is a public and trivial data to be found (19, 20). For the calculation of the parameters *N*_*sat*_, *q and a*, of equation (7), we use the model presented in (19, 20), where:

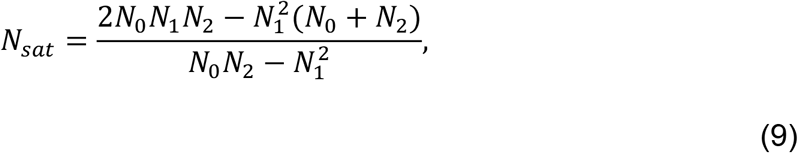

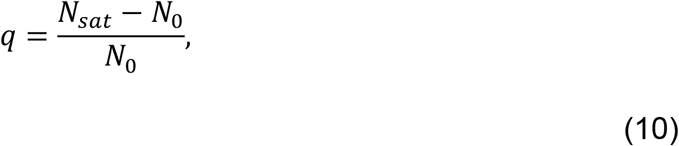

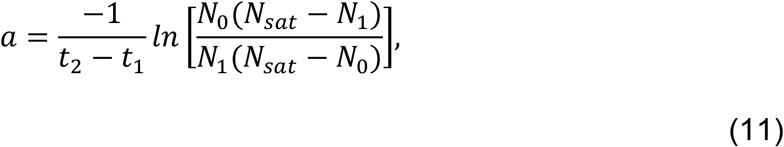

here *N*_0_, *N*_1_*eN*_2_ are constants and equidistant in time scale (19, 20). All data used to model this work were taken from the referenced sites (26 - 28).

## Discussion

A numerical analysis of the problem was carried out based on the above analytical mathematical model where the equation parameters are obtained using the approach described in references (7) - (11). We consider the cities of Niterói (Rio de Janeiro State) and Rio de Janeiro, but also Brazil, United States, and the world as a whole. The objective was to evaluate the scenario from micro to macro to try to understand in a more clear way the pattern of evolution of N(t). As time scale, we have considered the month of march as ground zero. Thus, in figures 1-5, on the abscissa axis, we have t = 0 = march (beginning of time) and t = 4 = July (final). However, the model allows extrapolations to be made for the months following July. Graphs of the population density of contaminated N(t) were produced over time and their derivatives, up to the third derivative, with the objective to studying the inflection at the growth rate of the curve and to check the critical points of *N*(*t*) and 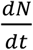 used to show inflections in the *N*(*t*) and 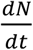 curves. Those critical points show us the time and the density at which inflections in the population and at the contamination rate contamination occur. This type of study clearly describes where and when the contamination rate changes, from increasing to decreasing, this via the first derivative of *N* (*t*).

**Figure 1.**
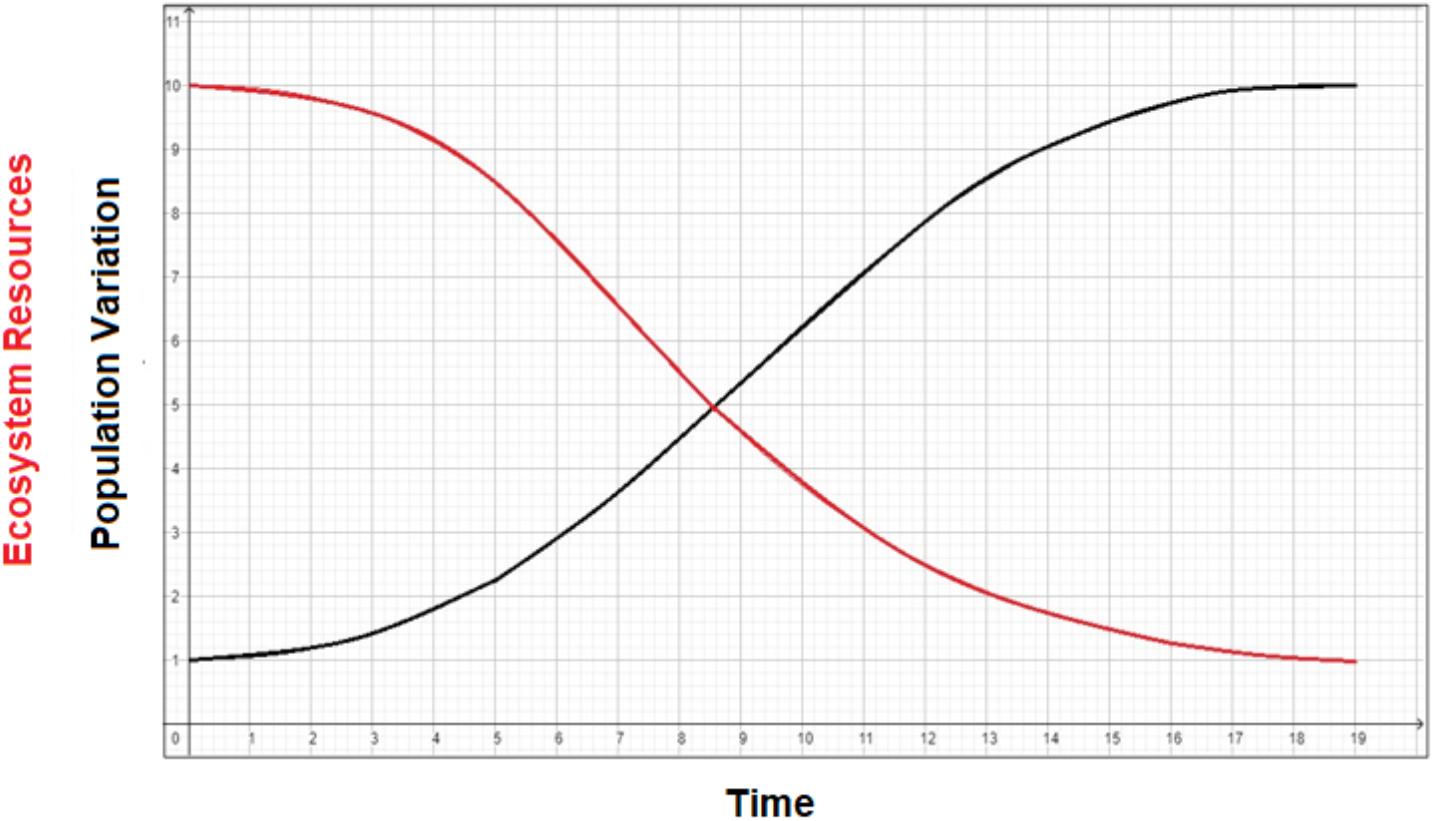
Scheme of population variation x Ecosystem resources.

The absolute numerical values shown in the graphs depend on the accuracy of the official information on the number of contaminated and on those who can be contaminated (population background of healthy people). In other words, our numbers may be underestimated by a factor (but not by orders), for this reason we created in our numerical model a data correction parameter when to adjust the parameters values is necessary. There is information in the media that this factor can be up to/higher than 12 times the reported values, however our simulations indicates that if this is the case, the number of contaminated people would be absurd, greater than the world contaminated number. According to the results calculated by the mathematical model used, a reasonable underreporting factor can be between 50% and 150%. In fact, simply multiply *N*_0_, *N*_1_*eN*_2_by a fiiting parameter, *SN*_0_, *SN*_1_*e SN*_2_, adjusting them for the “correct” notification, in which case *SN*_0_ =1, *SN*_1_ =1 *e SN*_2_ =1, or whatever *SN*_0_ =%*N*_0_, *SN*_1_ =%*N*_1_*e SN*_2_ =%*N*_2_.

Figures 2-6 show N(t) that describes the temporal evolution of the number of infected people, and their derivatives that describe the critical points of the curve are also presented. Figure 1 shows the cases of Rio de Janeiro city and Niterói city. For Rio de Janeiro city, the numbers foreseen is of the order of 100,000 contaminated people, at the saturation level, it seems that we are slowly going through the inflection of the curve (with some overshooting). The saturation for the case of Rio de Janeiro, predicted by the model is for July or even August if there is a negligence in the containment / contingency measures. If the real measure exceeds 100 thousand, there were certainly under-notifications for *N*_0_, *N*_1_*e N*_2_.

**Figure 2.**
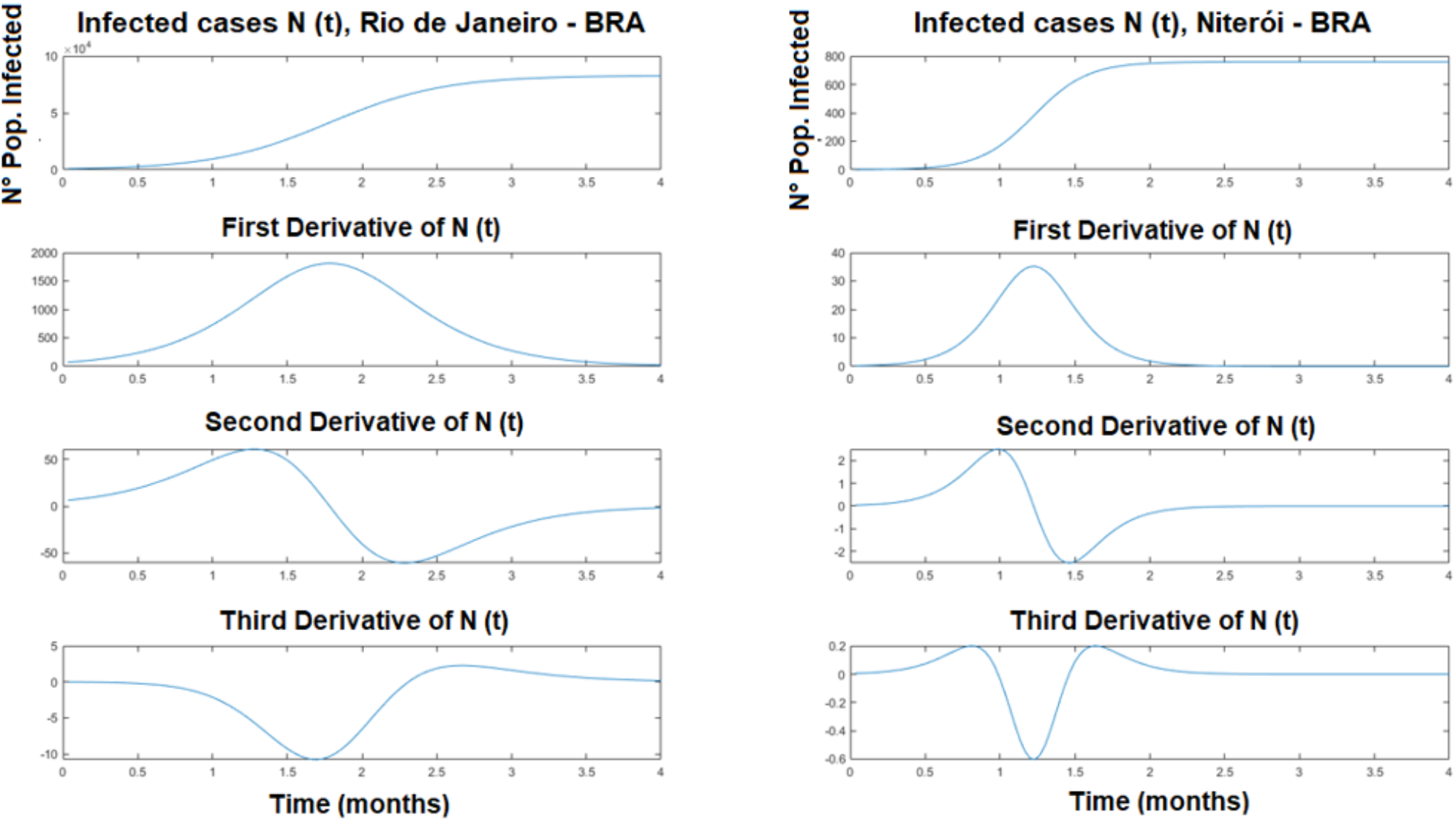
Population projection N(t) of infected cases and projections of saturation level for the cities of Rio de Janeiro and Niterói.

**Figure 3.**
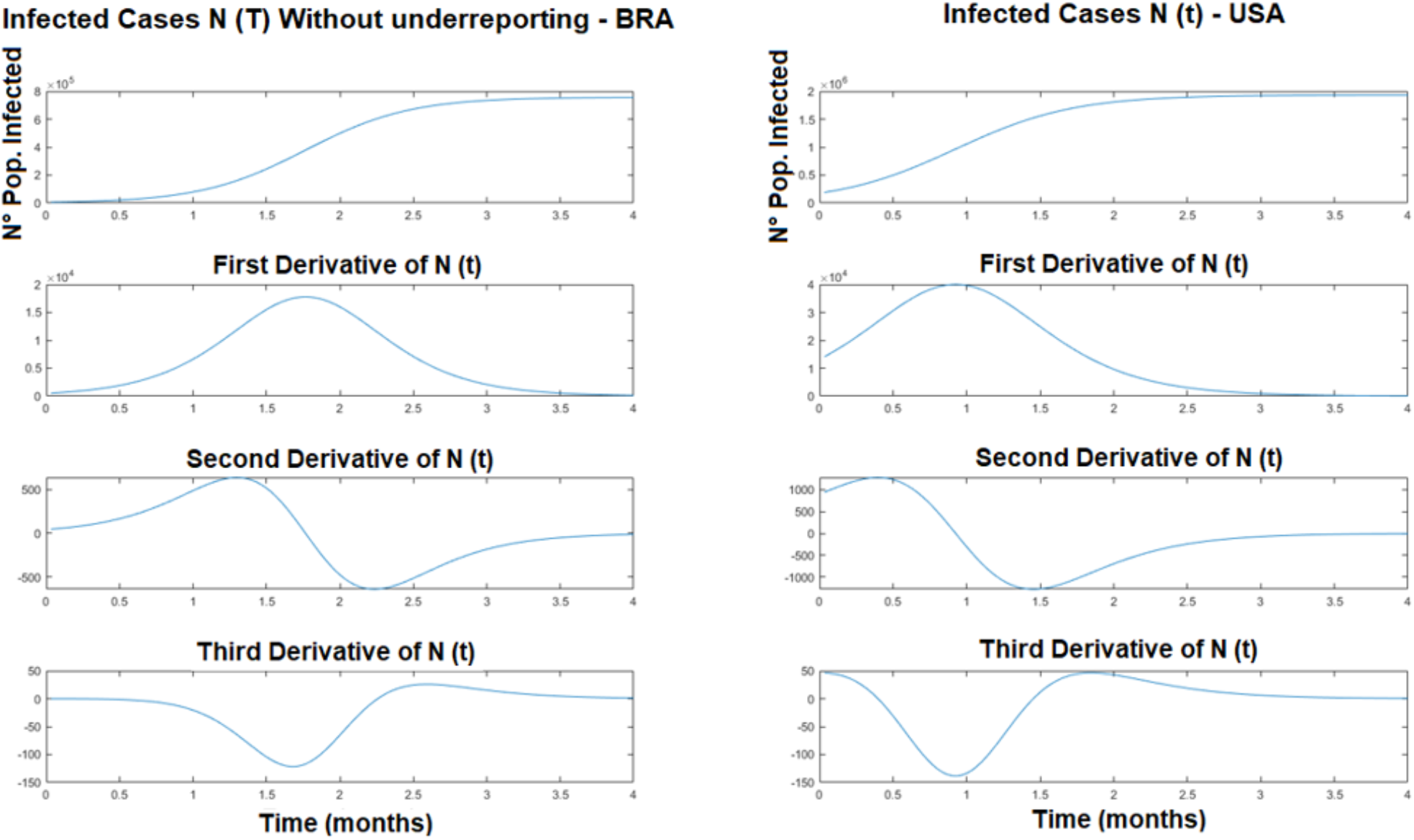
Projection cases infected population N(t) and the saturation level of Brazil projections (not underreported) and USA.

For Niterói, the simulation shows a plateau regime close to june, with 800 contaminated. However, the real number for the contaminated population is higher by a factor, indeed 4,327 contaminated in june 22 (covid.saude.gov.br). Later in july/august the real data will be even higher, by close an order of magnitude considering the value of our actual simulation. A simulation understimation by a factor (or by one order of magnitude) is very reasonable in this scenario, if we take into consideration that The OMS considers the data in Brazil underestimated, due to the low test rate. Health related personal consider that Brazil is testing much less than it should by at least one order of magnitude.

Since the rate of testing is improving with time, If we correct the simulation input data by one order of magnitude, the curve might be considered just fine at the real time of measure, for the logistic model used here.

The same discussion is valid for Rio de Janeiro city.

In Figures 2-6, the first derivatives show the exact moment when the slowdown of the contamination rate starts (after the curve maximum). The second derivatives show (at the zero value) the moment when the inflection of N(t) begins, and also the changes in concavity. The third derivatives show the exact moment of 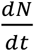 inflection and how its concavity changes with time. An inflection in 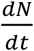 shows clearly the 3W (how, when, where) the contamination growth rate reaches its maximum value and how it changes in time evolution. The third derivative is used just to ensure that the maximum calculation/inflection of 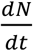 is indeed correct. Note that 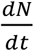 has two inflection points, one close to t = 1 and the other close to t = 2.2.

The concavity starts upwards, then inverts after the first inflection point and inverts again at the second inflection point, clearly showing that the contamination growth rate increases, reaches the maximum and then decreases with time. It is not possible yet to see this effect on N(t), this because this is not a linear effect. In other words, *N*(*t*)still seems saturated, but the contamination rates are on a downward trend, later afterwards this effect will indeed impact N(t) in a measurable way, and it will start to fall in time in a visible way.

Note that *N*(*t*)has an inflection point due to the nature of the logistics curve, but its first derivative, which models the rate of change of the population without time, has two inflection points, showing an increase and decrease in the rates. Also, due to the nature of the logistic model used here, we cannot model the decreasing phase of *N*(*t*), we just know it will happen naturally after saturation, if a second wave does not occur.

Figures 4 and 5 take into account possible cases of underreporting in the actual number of infected cases, where 2x and 12x, means that numbers were considered two and twelve times more than those reported by the Ministry of Health in Brazil.

**Figure 4.**
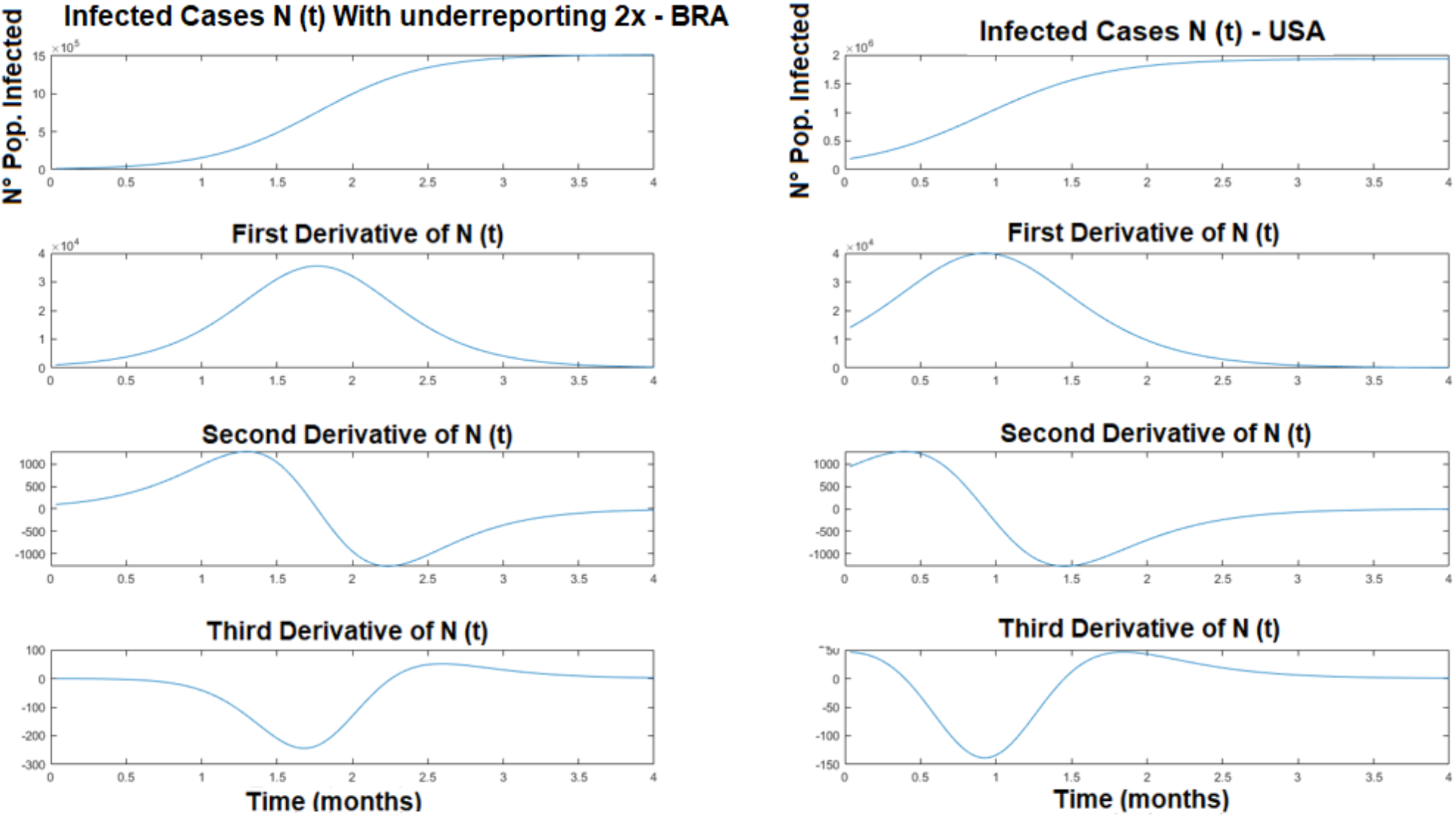
Population projection of infected cases N(t) and saturation level projections for Brazil (with underreporting of 2x more cases) and the USA.

**Figure 5.**
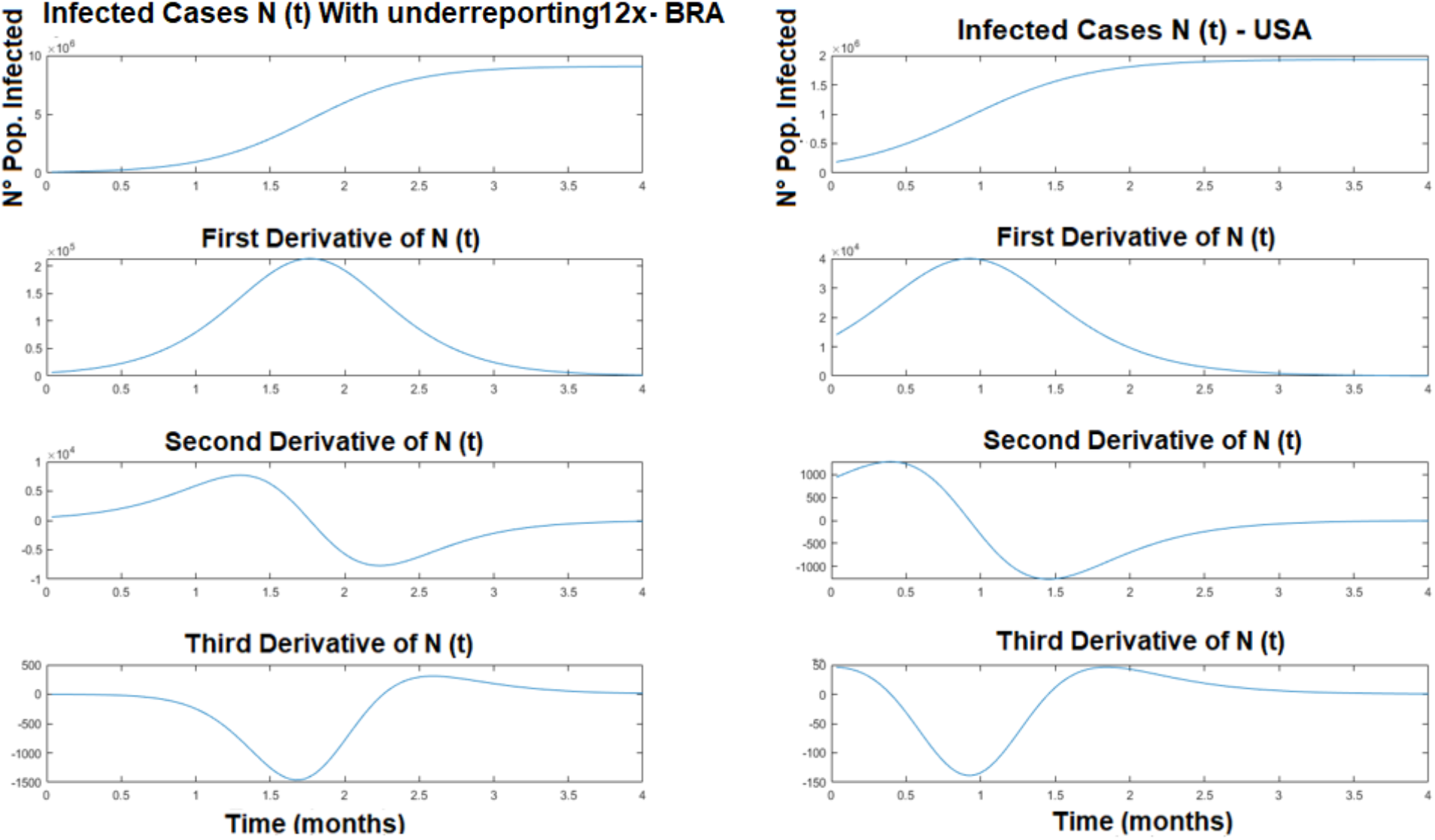
Population projection of infected cases N(t) and saturation level projections, for Brazil (with underreporting 12x more cases) and USA.

In Figure 6 shows, just for comparison, the number of cases of infected people in Brazil (without underreporting) and the number of cases in the World (Global).

**Figure 6.**
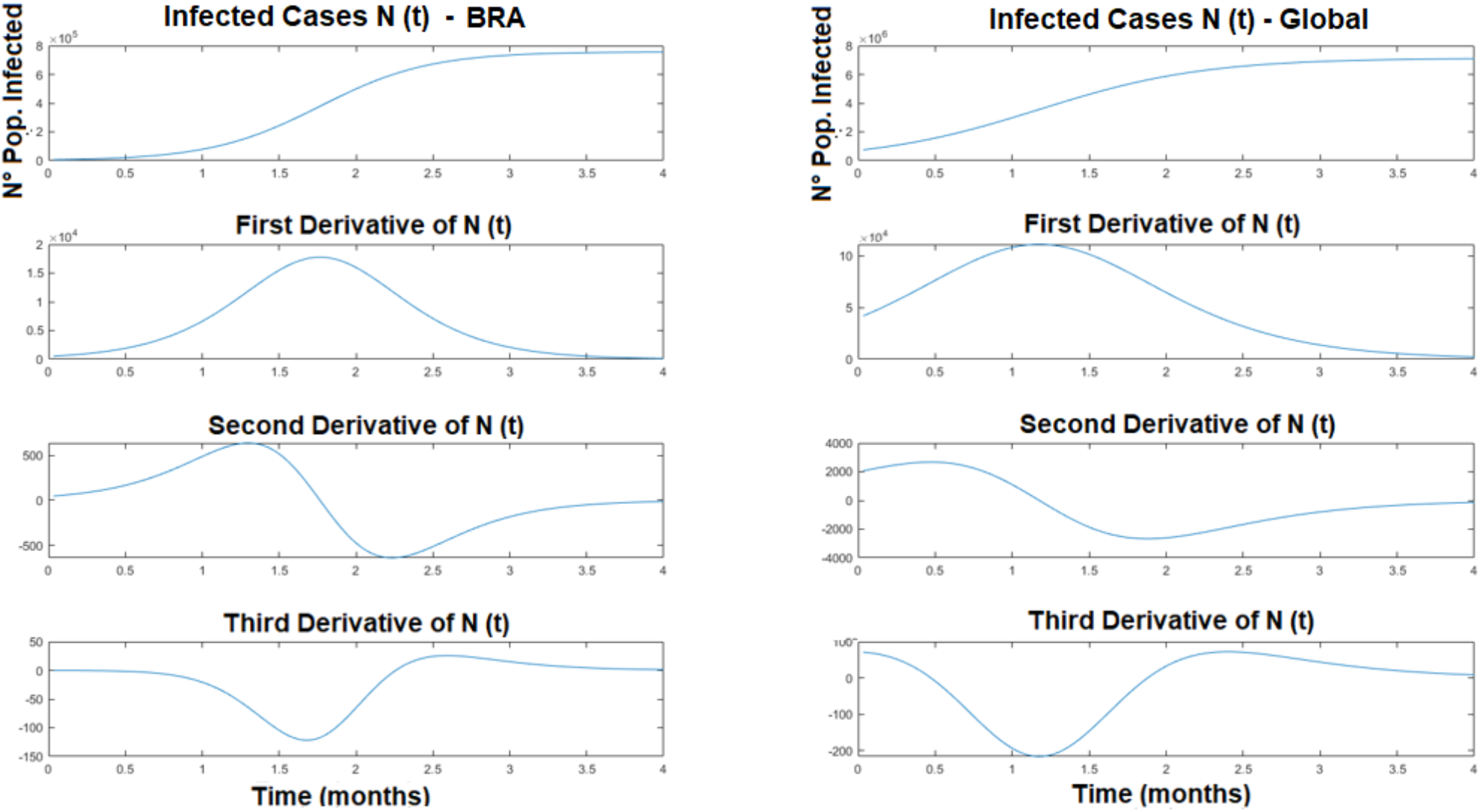
Population projection of N(t) infected cases and projections of saturation level, for Brazil (without underreporting) and Global.

## Conclusion

Based on the mathematical model proposed in this work, it can be seen that to reduce the number of contaminated people per unit of time, it is necessary that the factor *bN*(*t*), in equation 3, to be increased in time, this is only possible with the death of the population little by little, an effective vaccine, or with quarantine/social isolation. If the social isolation is done with prevention so that the aerial contamination is contained and the fomites as well. Just counting on the reduction of the amount of contaminated people per unit of time leaving the system free as the Swedish model of herd immunization, it seems that it has so far been inefficient, and has led to the death of a large number of infected people. At the moment, without a vaccine to contain the COVID-19, the most “antiviral” tool possible under our absolute control is not giving the virus the opportunity to reproduce itself with severe hygiene measures and social isolation. In the case of contamination, what remains is to seek specialized and competent medical help immediately, avoiding the transmission of the virus to other people and worsening its personal situation.

In this study, parameter b (impact parameter between elements of the target population with unit of measure 1/dim(*tN*(*t*)), measures how the reduction in the contaminated population impacts the contamination of new elements of the public. This parameter that for reasons of simplification was considered constant in time, in real life it also depends on time, it increases as the number of people to be infected decreases, that is, if *N* (*t*), decreases with time *b*(*t*), increases.

This study only considered one population, but we know that in fact there are more than one interacting group, we could consider for instance: non-contaminated population, non-isolated contaminated population, isolated contaminated population, and government contingency actions.

If we consider two populations where one is the predator, the correct model to use would be the Volterra predator - prey model. If there are limitations, the predator-prey with a limitation term applies, as in the logistical case (which is a model for just one population). If there are three interacting populations, then the dynastic model (super predator, predator, prey) must be used. In the case of predator-prey there is the possibility of ecological balance, this type of balance must be avoided at all costs, and this is the “least” nonlinear case possible for population dynamics of coupled populations (12).

To understand better who the populations would be in those cases, it can be said that in the dynamics of the predator-prey, one can consider government actions (or number of contaminants that the government effectively serves and leaves the system - via hospitals, doctors, nurses) and the number of contaminated. If the government does not apply serious and competent policies, this system can go into an ecological balance, that is, if the number of contaminated people falls, the government relaxes, the number of contaminated people grows, and so the government intensifies again its actions, and for this reason the number of contaminants decreases again, the government relaxes again, and so it goes over time forever, this is a true ecological balance between of the government actions and the number of contaminated people (and us). In the dynastic case, there are three populations, the super predator, is the government, the predator might be the uncontaminated society, and the “prey” the contaminated subjects, but depending on what one wants to study, the predator population can change the position to be prey and vice versa, but in any case, chaos and bifurcations can occur in this type of population dynamics, as in the climate, whose forecast depends dramatically on the initial conditions (21). For this reason, caution should be at hand when to make predictions about how the society will be like at the end of this pandemic time (21-23).

Many effects “foreign” to the expected normality occur in the population dynamics, such as the so-called “population irruption” or a sudden and impetuous action of elements of the population in such a way that it exceeds (“overshoots”) the load capacity of the system, causing a change in this capacity and forcing the system to stabilize at another level, if not before causing irreversible or long-term damage to the ecosystem in which this population is inserted. The population can also, after the “overshooting”, present damped and non-damped oscillations in the vicinity of the system’s carrying capacity.

In other words, the number of contaminated people can fluctuate in the region of the carrying capacity, depending on the environment in which the population is inserted. In this case, it would be an anomalous contamination to the carrying capacity of the healthy population to sustain the action and spread of the virus.

Mathematical modeling indicates that the greater the social isolation, the reduced the non-linearities in the system. The primary role of the government, if there is a drastic reduction in the circulation of people, is to have the competence to lead the country’s economy, avoiding the economic collapse, we know that the world system is closed and there is always “self-organization” in every non-linear system, the world economic system as such will certainly have a “self-organization” and the flow of capital and goods will take its course, but it will continue to flow as in the past, maybe at a different path. If there is social isolation for those who have no real need to move, those who have this need may keep part of the economy spinning but with physical and biological security, so as not to fall ill when sacrificing for others (24, 25).

Finally, it is worth mentioning that there are numerous methods used to study the dynamics of population growth, stabilization, and reduction / death. Some of these methods are: a - arithmetic model, b - geometric / exponential model, c - multiplicative regression, d - decreasing growth rate, in addition to the logistics studied in this work. The choice of the logistic model is due to the fact that the arithmetic, exponential, and geometric growth rates are not limited by the saturation density (*N*_*sat*_) obtained with the logistic model. The geometric, exponential, and linear rates approximate the problem reasonably well until the inflection or a little earlier, but they depart from the logistic rate close to the inflection region of the inflection, therefore these methods provide incorrect results after inflection and do not show saturation, as *N*_*sat*_ obtained with the logistic model, which is the simplest model that describes the problem in a very acceptable way (19).

## Data Availability

PUBLIC DATA

https://covid.saude.gov.br/

https://www.who.int/

http://www.saude.niteroi.rj.gov.br/

## References

1. Parametric identification and public health measures influence on the covid-19 epidemic evolution in brazil, r.m. Cotta1, 2, c.p. Naveira-cotta2, and p. Magal, medrxiv preprint doi: https://doi.org/10.1101/2020.03.31.20049130 |xthis version posted may 12, 2020.

2. M. Batista, estimation of the final size of the covid-19 epidemic, medrxiv, (2020) 2020. 2002.2016.20023606.

3. Modelo logístico – brasil – covid 19 – 2020, observatório covid-19 Maringá, http://complex.pfi.uem.br/covid/.

4. Kriston l. Projection of cumulative coronavirus disease 2019 (covid-19) case growth with a hierarchical logistic model. [preprint]. Bull world health organ. E-pub: 7 april 2020. Doi: http://dx.doi.org/10.2471/blt.20.257386.

5. Gaussian temporal evolution from corona virus from the first case to the peak: the temporal average constant k, international journal on engineering, science and technology, volume 2, issue 1, 2020, antonio. J. Balloni, campinas/sp, rogério winter.

6. Situation report - 144, coronavirus disease 2019 (covid-19) 12 june 2020.

7. ‘Hubs of Infection’: How covid-19 spread through latin america’s markets, https://www.theguardian.com/world/2020/may/17/coronavirus-latin-america-markets-mexico-brazil-peru.

8. 8 - Johns Hopkins University of Medicine – coronavirus resource center <https://coronavirus.jhu.edu/map.html>.

9. Boyce, W.E.; Diprima, r.C. Elementary differential equations and boundary value problems. 7a. Ed. New york: john wiley & sons, inc., 2001.

10. http://coronavirus.butantan.gov.br/ultimas-noticias/o-que-e-imunidade-de-rebanho.

11. Verhulst, P.E. Recherches mathématiques sur la loi d’accroissement de la population. Académie de Bruxelles, Bruxelles, 18:1–38.

12. Harold T. Davis, Introduction to Nonlinear Differencial Equation, Dover, Nem York, 1960.

13. Estudo analítico da equação de Fisher linearizada: determinação de tamanhos mínimos de fragmentos populacionais / Renato Pacheco Villar, Dissertação de mestrado, universidade federal de alfenas, 2014.

14. Logistic models with time-dependent coefficients and some of their applications, raquel m. Lopez, benjamin r. Morin, and sergei k. Suslov, cornell university, 2010.

15. https://www.healthknowledge.org.uk/public-health-textbook/research-methods/1a-epidemiology/epidemic-theory.

16. https://www1.health.gov.au/internet/publications/publishing.nsf/Content/mathematical-models~mathematical-modelsmodels.htm~mathematical-models-2.2.htm].

17. The Precautionary Principle Also Applies to Public Health Actions Bernard D. Goldstein, Am J Public Health. 2001 September; 91(9): 1358–1361.

18. ALARA - As Low As Reasonably Achievable, https://www.cdc.gov/nceh/radiation/alara.html, USA Certers for Disease Control and Prevention – CDC.

19. Wastewater Treatment Plants: Planning, Design, and Operation, Second Edition, Syed R. Qasim, Routledge, 1985 - Technology & Engineering.

20. Von Sperling M. (2014). Princípios do tratamento bioógico deáguas residuárias. Vol. 1. Introdução à qualidade das águas e ao tratamento de esgotos. Editora UFMG. 4a ed., 472 p.

21. The Dynastic Cycle and the Stationary State, Dan Usher, The American Economic Review, Vol. 79, No. 5 (Dec., 1989), pp. 1031–1044.

22. N. Fiedler – Ferrara C. P. e C. P. Cintra do Padro, Caos: Uma introdução, Edgar Blucher, 1994.

23. Generalized logistic growth modeling of the COVID-19 outbreak in 29 provinces in China and in the rest of the world Ke Wu, Didier Darcet, Qian Wang and Didier Sornette, arXiv:2003.05681 [q-bio.PE], Cornnell University, 2020.

24. Social Isolation in Modern Society, R. Hortulanus, Anja Machielse, and L. Meeuwesen, Routledge, Taylor & Francis Group, London, 2006.

25. Social Isolation, Loneliness, and All-Cause Mortality in Patients With Cardiovascular Disease: A 10-Year Follow-up Study, Yu Bin, Andrew Steptoe, Li – Jung Chen, and Po – Wen Ku, Psychosomatic Medicine, 2019.

26. https://saude.gov.br/component/tags/tag/oms

27. http://www.saude.niteroi.rj.gov.br/

28. https://www.who.int/emergencies/diseases/novel-coronavirus2019?gclid=CjwKCAjw57b3BRBlEiwA1ImytjOFiJH189_Ax1it4CZJmFB3paFsewKw8jnW5aqHbOgdWiNJP2f9hoCuSQQAvD_BwE

